# Critically Ill Children Frequently Receive Medications with Established but Unused Pharmacogenomic Guidelines: Actionable Findings from an Integrated Electronic Medical Record and Exome Sequencing Study

**DOI:** 10.64898/2026.07.16.26358240

**Authors:** Noelle Lynch, Naama Elefant, Anya Revah-Politi, Andrew S. Geneslaw, Jaimee Beckett, J. B. Wall, Carlos A. Breton, Maya Sabatello, Steve Kernie, Hulya Bayir, Ali G. Gharavi, Joshua E. Motelow

## Abstract

**Importance:** Pharmacogenomic (PGx) guidelines can improve medication efficacy and reduce toxicity, but their application in pediatric intensive care units (PICUs) remains largely unexplored.

**Objective:** To determine the frequency of medications with established PGx guidelines administered in the PICU and assess the capacity of exome sequencing to capture PGx phenotypes for these medications.

**Design:** Retrospective cohort study integrating electronic medical record and exome sequencing data.

**Setting:** Morgan Stanley Children’s Hospital of NewYork-Presbyterian, a single center tertiary care children’s hospital.

**Participants:** A total of 4,939 children admitted to the PICU (2020 - 2024), and 192 children admitted to the PICU who underwent exome sequencing for research purposes (2015 – 2023).

**Exposure:** Critical illness requiring PICU admission.

**Main Outcomes and Measures:** Frequencies of administration of medications with established PGx guidelines in the PICU and the proportion of individuals with exome sequencing with identifiable PGx phenotypes.

**Results:** Among 4,939 PICU patients, 37.2% (n=1,837) received at least one medication with established PGx guidelines and 14.4% (n=712) received two or more such medications. Twenty PGx genes were implicated; *CYP2C9* was most common (17.3%, n=853). An estimated 8.2% of patients received medications for which PGx-guided recommendations would have altered clinical management. Among 192 patients who underwent exome sequencing, at least one metabolizer phenotype was identified in 62% (n=119).

**Conclusions and Relevance:** Many critically ill children receive medications with established PGx guidelines. This study highlights an opportunity for more personalized medicine for critically ill children admitted to a tertiary care hospital and assesses the strengths and weaknesses of exome sequencing to uncover pertinent PGx phenotypes.

**Key Points:** *Question:* Do critically ill children in a tertiary care hospital receive medications for which PGx guidelines exist, and does exome sequencing reliably identify relevant pharmacogenomic phenotype?

*Findings:* Analysis of electronic medical records from 4,939 critically ill children and exome sequencing of 192 patients demonstrated that many received medications that have established pharmacogenomic guidelines, and exome sequencing identified actionable pharmacogenomic variants with implications for treatment.

*Meaning:* Many critically ill children receive medications for which PGx guidelines exist but are not routinely applied. This represents a missed opportunity to apply precision medicine to improve the care of critically ill children.

## Introduction

Although Pharmacogenomics (PGx) guidelines exist for many commonly used medications, their application in the PICU remains limited. PGx refers to how genetic makeup affects medication response.^1–3^ PGx testing and associated practice guidelines aim to optimize drug efficacy and avoid adverse drug events (ADEs) by identifying patients who require deviations in standard dosing or the use of an alternative drug due to germline genetic variation. PGx testing implementation in pediatrics has identified successes and barriers.^4–13^ Beyond pediatrics, implementation in several clinical settings has been explored, including for individuals with critical illness.^14–17^ Patients in the PICU are at high risk of ADEs due to narrow therapeutic windows, polypharmacy, and increased sensitivity to the consequences of ADEs or reduced drug efficacy.^18,19^ PGx testing could significantly impact PICU patients, yet this has not been tested. Identifying the overlap of actionable PGx phenotypes with medications administered in the PICU would support feasibility of PGx-guided medication administration in the PICU.

Exome sequencing (ES) is an effective tool in the PICU for diagnosis and management.^20–26^ While ES is limited to gene coding regions and does not effectively identify structural variants, it can identify PGx phenotypes for many genes.^26–28^ Given the efficacy of ES in diagnostic testing, understanding its efficacy in detecting PGx phenotypes would allow for additional clinically beneficial applications.^27^

## Methods

### Study Population

We studied two populations: (1) “EMR-only”: We examined retrospective medication administration records (MAR) from the electronic medical record (EMR) of 4,939 PICU patients to identify medications with PGx recommendations that were administered. Patients were included if they were admitted to the PICU or cardiac intensive care unit (CICU) at New York-Presbyterian Morgan Stanley Children’s Hospital (MSCH)/Columbia University Irving Medical Center (CUIMC) and administered ≥ 1 medication in the PICU/CICU between 2/1/2020 and 2/1/2024. Medications administered to patients prior to their 18^th^ birthday were counted.

Genotypes were not available for this cohort. The Columbia University Institutional Review Board approved the study protocols (AAAV0962). (2) “EMR+ES”: We examined a dataset including both EMR and exome sequencing (ES) data. Children with critical illness were enrolled at MSCH/CUIMC (n=192) according to previously-described methodology.^23,29^ Critical illness was defined by admission to the MSCH/CUIMC PICU/CICU and administration of at least one medication. Written informed consent was provided for using DNA in genetic research was provided by the legal guardian for participants younger than 18 years or by the individual when 18 years or older. Exomes were collected from 7/23/2015 to 12/14/2020 (collection dates for six samples were unavailable). EMR data was collected from 5/24/2001 to 3/7/2023. Exomes were analyzed retrospectively, and pharmacogenomic phenotypes were not incorporated at the time of medication administration. The Columbia University Institutional Review Board approved the study protocols (AAAO8410, AAAO7952).

### Identification of medications administered using the EMR

The EMRs from MSCH span the use of two different EMR systems. EMR1 was prior to 2/1/2020 and EMR2 was active from 2/1/2020 onward. For the EMR cohort, all data were drawn from EMR2. For the EMR+ES cohort, the data were drawn from both EMRs. Medications administered (as opposed to ordered in EMR but never received) were identified using administration codes (eTable 1). MAR for the EMR-only cohort was collected from EMR2 and data for the EMR+ES cohort was collected from EMR1 and EMR2.

### Pharmacogenomic guidelines for EMR-only cohort

Guideline data on medications and phenotypes were downloaded from The PGx Knowledgebase (PharmGKB) (downloaded 2/4/2025).^30^ PharmGKB contains annotations on 105 unique medications (eTable 2) with annotations from either Clinical PGx Implementation Consortium (CPIC)(87), the Dutch Pharmacogenetics Working Group (DPWG)(53) or both.^31,32^ Of these medications, 63 contain a recommendation for a dose adjustment, 87 contain a recommendation for an alternative medication, 56 contain a recommendation pertaining to neither dosing nor alternative medications, and 52 contain information about the use of PGx tests in the context of prescribing the medication.^33–36^ There are CPIC or DPWG recommendations pertaining to genotypes in 22 genes (eTable 3).

PharmGKB annotates 422 medications by the quality of their FDA label. We included clinically actionable label annotations including “Testing required”, “Testing recommended”, or “Actionable PGx” (213 medications)(eTable 4). We excluded medications with “Informative PGx”, “No Clinical PGx”, and “Criteria Not Met” because these labels did not indicate clinical implications.

Combining medications with CPIC, DPWG, or clinically actionable FDA guidelines, we matched 272 medications (eTable 5) to medication formulations in the EMR-only cohort.

### Matching medication formulations with pharmacogenomic medications for EMR-only

EMR medication formulations include additional information and cannot be directly matched to PGx guidelines. A formulation may contain the medication, dosage, and route among other information. For example, a warfarin formulation may be “WARFARIN SODIUM 5 MG OR TABS” or “WARFARIN ORAL SUSPENSION 1MG/ML” among other possibilities. To identify PGx medications, we matched multiple formulations to a single PharmGKB PGx medication name (e.g. warfarin) based on the presence of the medication within the formulation. If an EMR medication formulation contains a PGx medication name, the EMR formulation matched to the PGx medication. Matched (eTable 6) and unmatched (eTable 7) formulations were reviewed manually for accuracy. Formulations not administered systemically were identified with the following phrases (EX OINT, EX CREA, EX PATCH, EX GEL, OP OINT, OP SOLN, OP SUSP. IN NEBU, Oint, topical oint, Ophth Soln, Opht Soln, nebulizer solution, bladder irrigation, OPH OIN, Amikacin Sulfate Approval, INH SOL, lock 5mL) and excluded from matching. Prilocaine was excluded from both cohorts as it was disproportionately present in EMR1 over EMR2 (data not shown).

### Estimation of extreme PGx phenotype-medication exposure

To capture the impact of the most extreme PGx-derived information on prescribing implications, we estimated the frequency of the most “extreme” phenotype for medications administered ≥50 patients. We defined “extreme” as the phenotype indicating the most significant change in medication administration per the associated PGx guidelines (eTable 8). We excluded medications for which guidelines were generated based on high-risk HLA alleles. To estimate the number of individuals affected by at least one medication-extreme phenotype pair, we assumed that administration of each medication and the metabolizer/medication phenotype were independent events. We then calculated the following probabilities:

For each medication i:

P(received medication i AND has extreme PGx phenotype i) = P(medication i) × P(extreme PGx phenotype i)

Assuming independence across medications, the probability that a patient has no such events:

P(none) = Π(1-P_i)

Therefore:

P(receiving at least one medication while having an extreme phenotype) = 1-Π (1-P_i)

This is the probability that a patient receives ≥1 medication for which their PGx genotype predicts a clinically significant deviation from standard care.

### Exome sequencing and alignment

ES was performed at or transferred to the IGM. Established protocols have been used at the IGM to generate ES for more than 120,000 individuals.^37^ Exomes were aligned to human reference GRCh38 using Burrows-Wheeler Aligner Maximal Exact Match (BWA-MEM) or the Illumina Dragen aligner.^38,39^ Multiple capture kits were used (eTable 9).

### Variant calling, star allele, pharmacogenomic phenotype identification

Star alleles and PGx phenotypes were generated using PGx Clinical Annotation Tool (PharmCAT, v2.15.5).^40^ PharmCAT will attempt to generate star alleles and phenotypes for 18 genes (eTable 10) and corresponding phenotypes (eTable 11) and can issue guidelines for 182 medications (eTable 12). Four additional genes (*CYP2D6*, *HLA-A*, *HLA-B*, *MT-RNR1*) require outside calls and were not attempted due to the coverage limitations of exome sequencing data.^41^

We used Genome Analysis Toolkit (GATK - Broad Institute, Boston, MA, USA) HaplotypeCaller (v4.6.1.0) to generate the variant call file (VCF) for PharmCAT using GATK Best Practices with the pharmCAT positions as input for –allele, --max-mnp-distance 1, --output-mode EMIT_ALL_ACTIVE_SITES.^42^

Using the VCF file as input, PharmCAT generated star alleles and PGx phenotypes. Each named allele in PharmCAT was given a score based on the number of variant positions used to define the allele. If required positions were missing in the input VCF, PGx phenotypes for that gene/medication/subject combination were considered “partially covered”. Only gene/medication/subject phenotypes in which all required positions were called were considered “fully covered.”^43^ PharmCAT assigns a PGx phenotype for fully covered cases and, when possible, partially covered cases. It generates recommendations based on CPIC guideline annotations, DPWG guideline annotations, FDA label annotations, and FDA PGx associations.^26,27,38^

### Matching medication formulations with pharmacogenomic medication and guidelines for EMR+ES cohort

Pharmacogenomic guidelines for the EMR+ES cohort were based on recommendations from PharmCAT for pharmacogenomic phenotypes derived from ES. We define ‘clinically actionable PGx guidelines’ as recommendations supported by CPIC, DPWG, or FDA labeling. There are 156 medications (eTable 13) in both PharmCAT and the clinically actionable list from the EMR-only cohort. For these medications, matching was done in the same manner as in the EMR-only cohort to create matched (eTable 14) and unmatched (eTable 15) formulations. The following 26 medications for which PharmCAT issues guidelines were excluded because they do not have “clinically actionable” guidelines (see Pharmacogenomic guidelines for EMR-only cohort): avatrombopag, carvedilol, darifenacin, diazepam, dolutegravir, donepezil, elagolix, esomeprazole, fesoterodine, flibanserin, galantamine, mavacamten, metoclopramide, mirabegron, nateglinide, nebivolol, pazopanib, perphenazine, propranolol, protriptyline, rabeprazole, raltegravir, tolterodine, and viloxazine. Conversely, there are 116 medications (eTable 16) in the clinically actionable list from the EMR-only cohort for which PharmCAT does not offer recommendations.

### Data analysis

Data were analyzed and visualized using R (v.4.3.1).^44^

## Results

### EMR-only cohort: medications with existing PGx guidelines administered to children with critical illness

The EMR-only cohort (Figure 1) included 4,939 patients who were admitted to the pediatric or cardiac intensive care unit (PICU/CICU) and administered at least one medication, over the years 2020-2024. For these patients, 118,136 medication formulations were retrieved from their EMRs of which 5,081 (4.3%) were matched to 70 unique medications with PGx guidelines (eFigure 1, eTables 6-7). 37.2% of patients (1,837/4,939) were administered at least one medication with existing PGx guidelines, and 14.4% (712/4,939) were administered two or more (Figure 2, eTable 17). The most administered medications with PGx guidelines were ibuprofen (14.8%), pantoprazole (9.3%), hydralazine (6.0%), ondansetron (6.0%), tacrolimus (3.7%), clobazam (2.7%) and warfarin (2.2%) (Table 1, eTable 18).

**Figure 1:**
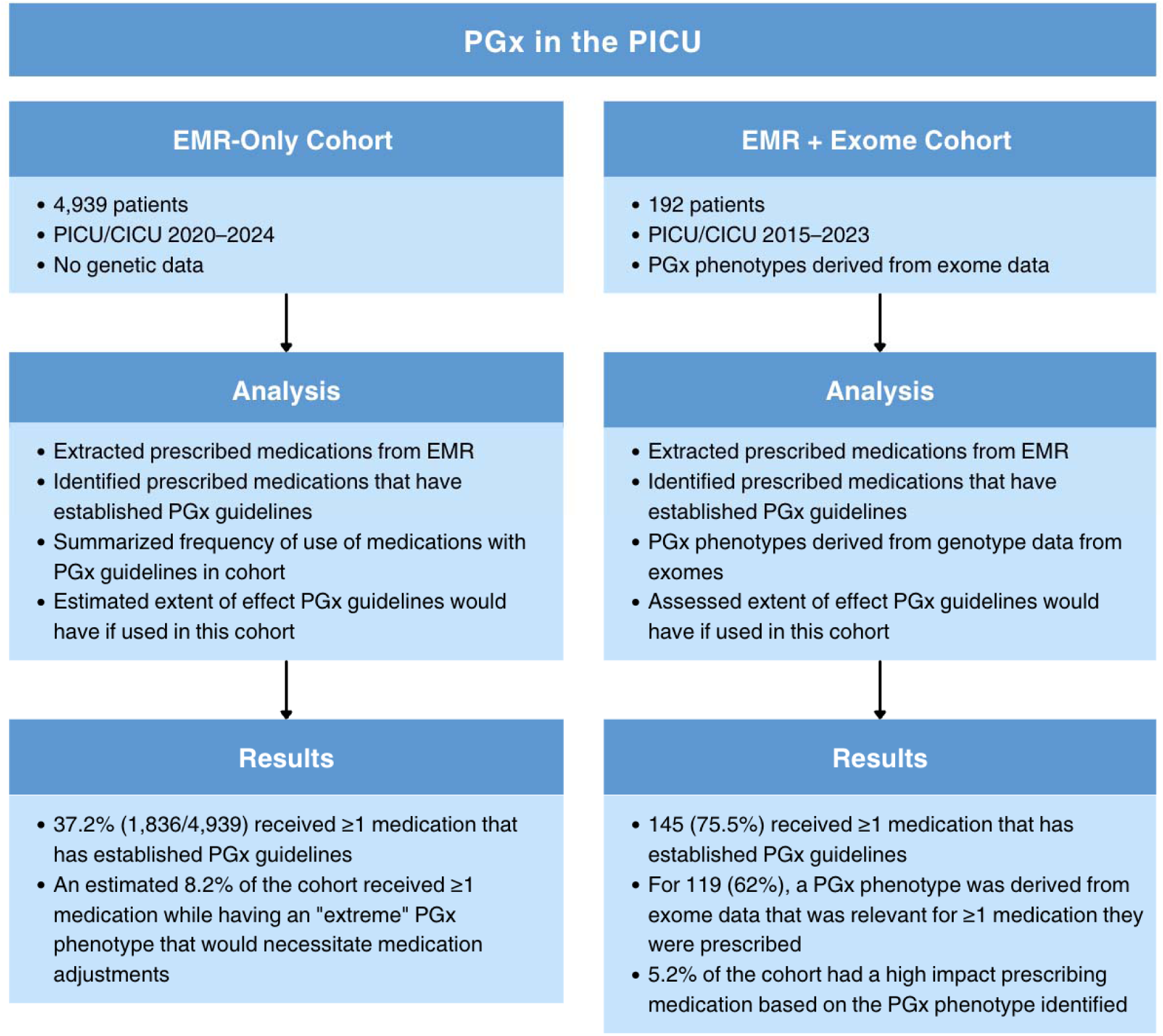
Study overview. Schematic of the study design showing the two cohorts analyzed. The EMR-only cohort (n=4,939) was used to identify medications with established pharmacogenomic (PGx) guidelines administered in the PICU. The EMR+ES cohort (n=192) integrated EMR data with exome sequencing to identify PGx phenotypes and assess exome coverage of PGx phenotypes and potential genotype-guided prescribing recommendations for administered medications. PGx guidelines were defined based on CPIC, DPWG, or FDA annotations.

**Figure 2:**
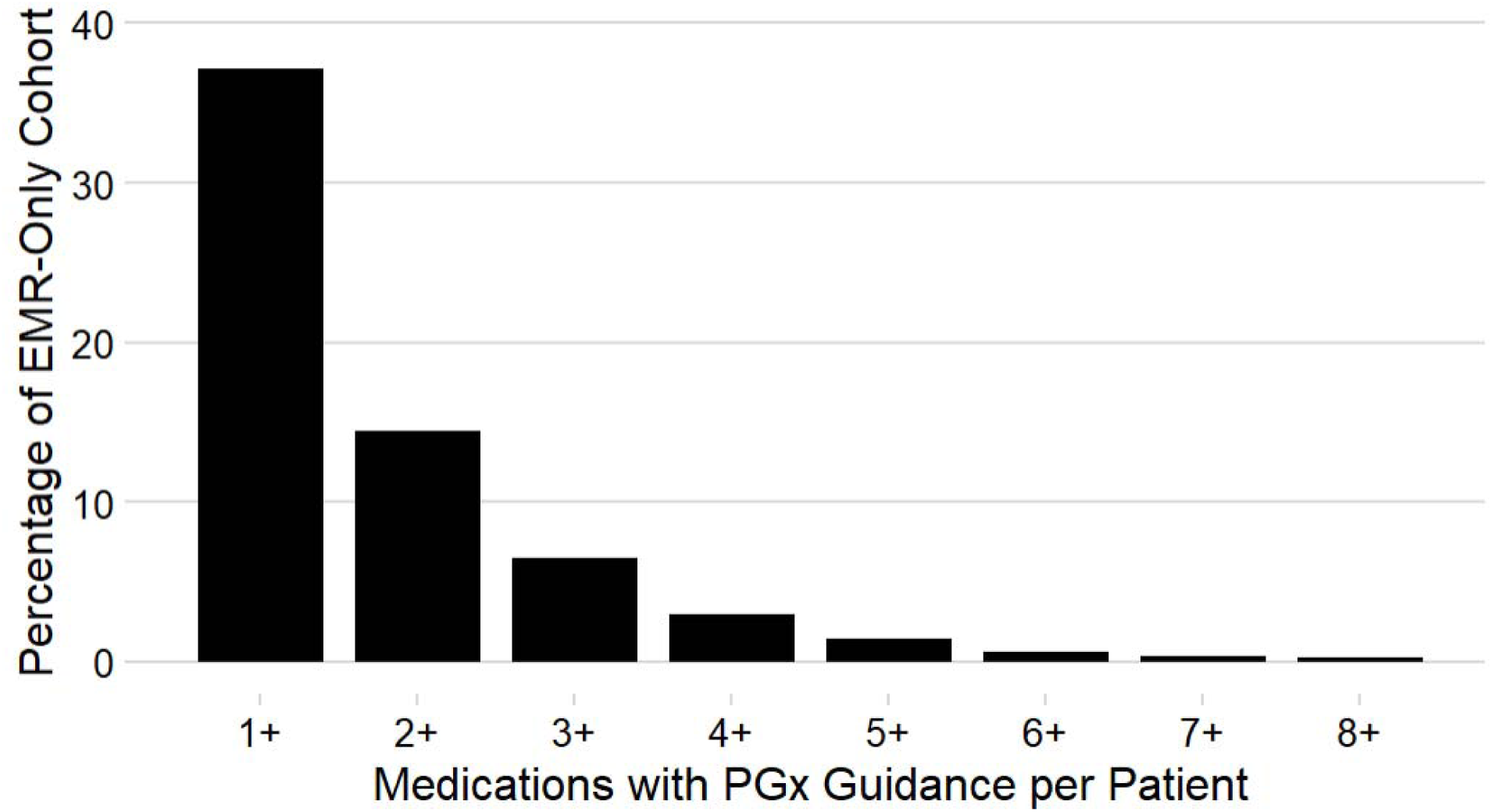
Pharmacogenomic medications per PICU patient in EMR-only cohort. Distribution of the number of medications with established PGx guidelines administered per patient in the EMR-only cohort. The x-axis shows the number of unique medications with established PGx guidelines administered per patient, and the y-axis shows the percentage of the cohort. 70 unique medications, range 0 to 12 medications with established PGx guidelines per patient, median=0 medications with PGx guidelines per PICU patient, interquartile range 0-1 medications with PGx guidelines per PICU patient. 3,102 patients administered 0 medications with PGx guidelines (not shown). PGx guidelines were defined based on CPIC, DPWG, or FDA annotations. PGx = pharmacogenomic, PICU = pediatric intensive care unit. Full data in eTable 17.

**Table 1:**
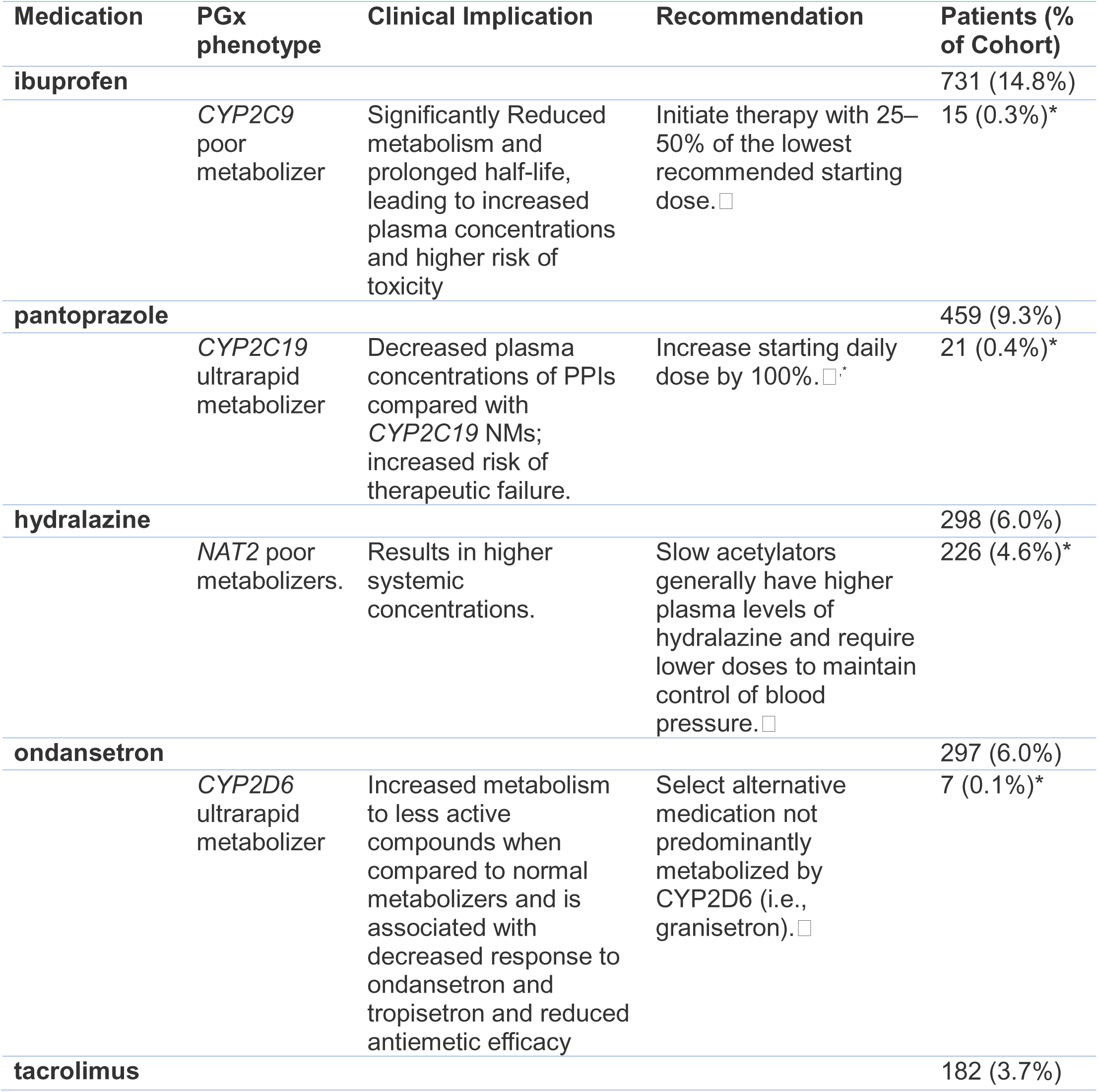
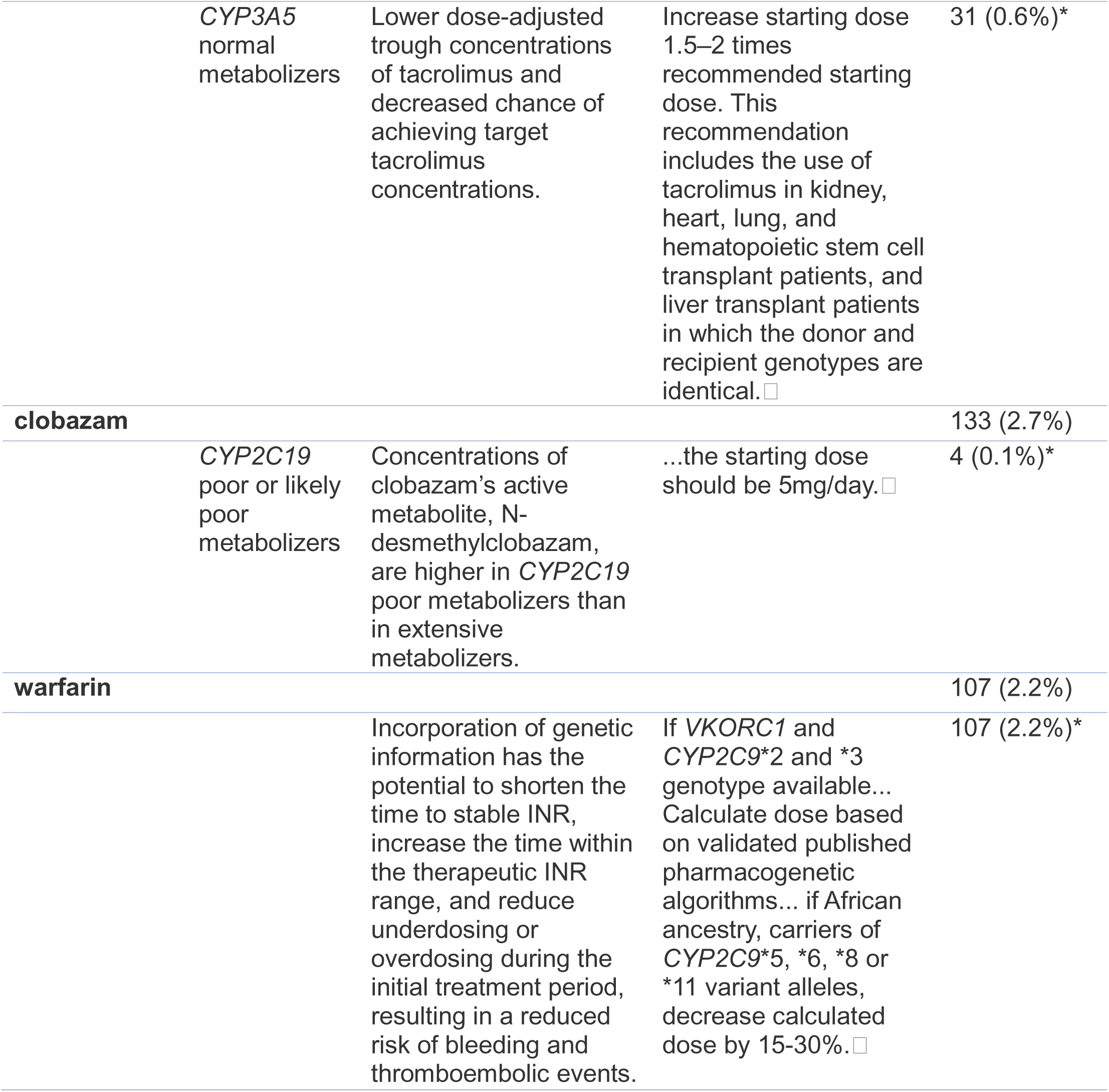
Number of children with critical illness taking specific medications with pharmacogenomic guidelines. Only medications administered to at least 50 patients are shown. ‘Phenotype’ column indicates the PGx phenotype with the largest change in clinical management. ‘Approximate affected patients (%)’ indicates the estimate of individuals in the total cohort who took the medication with the ‘Phenotype’. See eTable 5. ‘Implication’ indicates the effect of the phenotype on metabolism. ‘Recommendation’ indicates the CPIC or DPWG recommended change in clinical practice for the phenotype. Medications with HLA risk alleles (lamotrigine, oxcarbazepine) or PGx guidelines based on a co-condition were excluded (valproic acid). CPIC = Clinical Pharmacogenetics Implementation Consortium, DPWG = Dutch Pharmacogenomics Working Group, FDA = Food and Drug. Ll indicates CPIC guideline, Ll indicates DPWG guideline, Ll indicates FDA guideline, * indicates guidelines taken from FDA label directly and not generated by PharmCAT.

### EMR-only cohort: Estimated PGx phenotypes

While genotyping data were not available for this cohort, we estimated the prevalence of the clinically actionable PGx phenotypes based on known population frequencies, for all PGx-medication administered to at least 50 children (Table 1). We estimated that 0.3% of our cohort (∼15 individuals) likely received ibuprofen while having a PGx phenotype associated with reduced metabolism, which would result in high medication levels. These patients would have been at higher risk of acute kidney injury unbeknownst to the prescribing team.^45,46^ We estimate that pantoprazole was prescribed to 0.4% of our cohort (∼21 individuals) with standard prescribing guidelines despite having a PGx phenotype that would lead to decreased plasma concentrations. PGx established guidelines would recommend to double the starting daily dose for these patients.^47^ While over one-third of patients were exposed to PGx-relevant medications, we estimate that 8.2% of the cohort (∼405 individuals) would receive at least one medication while having an “extreme” phenotype for that medication that would have altered care.

### EMR-only cohort: Pharmacogenes relevant to administered medications

The most implicated pharmacogenes (Figure 3, eTable 19) included *CYP2C9* (853 individuals, 17.3% of cohort; e.g., ibuprofen, phenytoin, warfarin), *CYP2C19* (633 individuals, 12.8% of cohort; e.g., clopidogrel, pantoprazole/lansoprazole, voriconazole), *CYP2D6* (462 individuals, 9.4% of cohort; e.g., ondansetron), *CYP3A5* (3.7% of cohort; e.g., tacrolimus), and *HLA-B* (167 individuals, 3.4% of cohort; e.g., allopurinol, fosphenytoin, oxcarbazepine).

**Figure 3:**
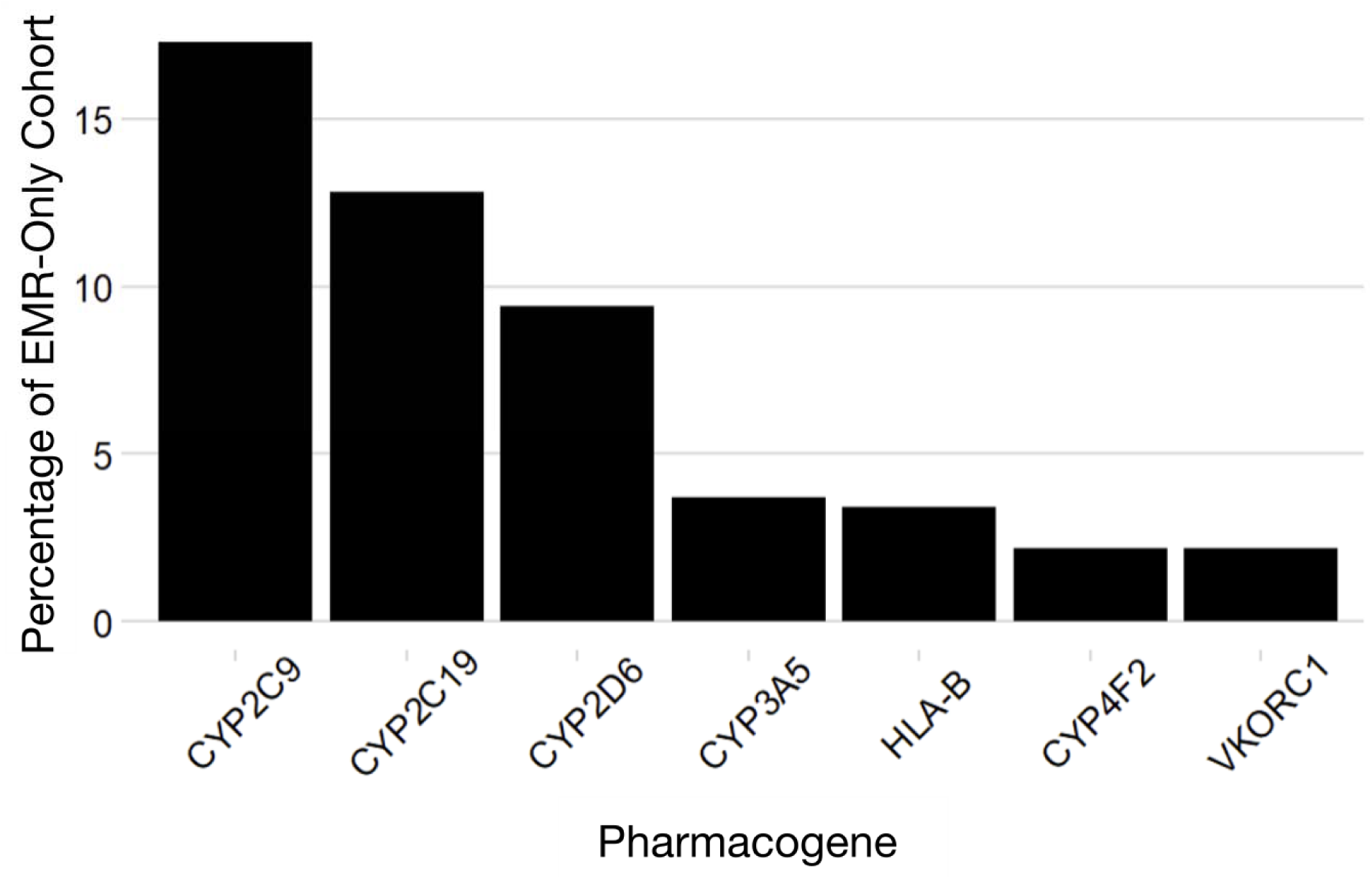
Pharmacogenes associated with medications administered to children with critical illness. Number of children receiving medications with established PGx guidelines, grouped by associated pharmacogene. The y-axis shows the number of patients, and the x-axis shows pharmacogenes linked to the administered medications. Only genes associated with medications administered to at least 90 patients are shown. A total of 4,939 patients and 22 genes were evaluated. PGx guidelines were defined based on PharmGKB annotations derived from CPIC, DPWG, and FDA sources.

### EMR+ES cohort: medications with existing PGx guidelines administered to children with critical illness

The EMR+ES cohort included 192 PICU/CICU patients (Figure 1). Similar to the EMR-only cohort, we pulled the medication formulations for these patients and found 10,784 unique patient and medication formulation combinations of which 608 (5.6%) were matched to 35 unique PGx medications (eFigure 3, eTables 14-15). We find that 75.5% (145/192) received at least one medication with existing PGx guidelines. The most commonly administered medication with PGx guidelines (eTable 20) was ondansetron (35.9%), followed by ibuprofen (29.2%) and pantoprazole (26.6%).

### EMR+ES cohort: Exome derived PGx phenotypes and Individualized recommendations

Using the available exome data for this cohort, we retrieved the patients’ star alleles and PGx phenotypes (see methods). While exome sequencing is not the ideal tool for identifying PGx metabolizer phenotypes, we evaluated ES not as a replacement for dedicated PGx testing, but as an opportunistic secondary use of existing genomic data. In this cohort, 70.3% (135/192) had adequate exome coverage to identify PGx genotypes and matching phenotypes for 40.0% (42/105) of the 105 medications investigated (eTable 2, 21-24).

Combining the medical formulation information from the EMR with the exome-derived PGx data (eFigure 4), we find that in 62% (119/192) of patients, PharmCAT provided PGx based recommendation for an administered medication. In 40.6% (78/192), the recommendations were based on a fully covered genotype and in 52.1% (100/192) they were based on a partially covered genotype and statistical assumptions (eFigure 4). An additional 34 individuals did not receive a phenotype due to inadequate coverage from ES.

We divided recommendations into low, medium, and high clinical impact (eFigure 5, eTable 25-26). 11 individuals had high clinical impact recommendations, 100 had medium and 103 had low clinical impact recommendations. Some examples of high clinical impact information are PGx metabolizer phenotypes that could have prevented prescribing clopidogrel to a likely non-responder, or prescribing tacrolimus and allopurinol at sub-optimal doses (see eTable 24). Most of the high clinical impact recommendations were based on fully covered positions (eFigure 5, 10/11 individuals).

## Discussion

Our study assesses the prevalence of the use of medications with established PGx guidelines in the PICU, the ability of exome sequencing to capture relevant PGx phenotypes, and the potential clinical implications when such data are available. While many commonly used medications have established PGx guidelines, such guidance is not routinely applied in clinical care. We found that medications with established PGx guidelines are administered widely in the PICU, with 37.2% of patients in the EMR-only cohort and 75.5% of patients in the EMR+ES cohort receiving at least one medication that has PGx guidelines, suggesting significant missed opportunities to personalize care (Figure 1). The higher proportion observed in the EMR+ES cohort (75.5%) likely reflects differences in patient recruitment, as children undergoing exome sequencing often represent a more medically complex group. These results align with previous studies on children with complex or chronic conditions which found that approximately 70% of these children had been prescribed at least one medication with PGx guidelines during their study period.^11,49^ Furthermore, previous studies on pediatric populations have found that approximately half of children carried variants in PGx genes for a current prescription or for a medication that was a relevant therapeutic option in their clinical management.^11,50^ In our study, the most commonly prescribed medication with established PGx guidelines was ibuprofen, which is associated with acute kidney injury (AKI) in hospitalized children, suggesting PGx informed dosing could modify the burden of AKI in the PICU.^51–53^ Despite this widespread exposure, PGx information is not currently integrated into prescribing decisions. We estimate that up to 8.2% of children in the PICU receive at least one medication for which established PGx guidelines would recommend a meaningful change in clinical management, underscoring a clinically significant missed opportunity to improve efficacy and reduce toxicity.

While ES is not the ideal modality for PGx testing, we evaluated whether existing ES data could help identify relevant PGx phenotypes in this population. We identified multiple medications administered in the PICU for which ES could fully identify the PGx phenotypes (Table 3). Examples included phenytoin/fosphenytoin and tacrolimus. In addition, ES identified several medications despite missing variants using statistical probabilities, including a case where clopidogrel was prescribed for a child with a PGx profile predicting minimal efficacy.

**Table 2:** Exome-derived pharmacogenomic (PGx) recommendations with explicit dose or therapy modifications. PGx phenotypes identified from exome sequencing are shown alongside associated clinical implications and recommended changes in therapy for medications administered in the EMR+ES cohort. Only recommendations with explicit dose adjustments or alternative therapy guidance are included. “Coverage” indicates whether all required genetic variants for phenotype assignment were fully captured by exome sequencing.

| **Medication**<br>**Gene**<br>**Phenotype** | **Implication** | **Recommendation** | **Covered(**<br>**Fully \|**<br>**not Fully)** | **Total**<br>**Patie**<br>**nts** |
| --- | --- | --- | --- | --- |
| **Allopurinol**<br>**ABCG2**<br>**Decreased**<br>**Function** | The effectiveness of allopurinol is reduced, meaning that a higher dose is required. The gene variation reduces the excretion of uric acid by kidneys and intestines, meaning that a stronger inhibition of the uric acid production by allopurinol is required to achieve the desired uric acid concentration. | Use 1.25 times the standard dose. | 1\|0 | 1 |
| **Clopidogrel**<br>**CYP2C19**<br>**Intermediate**<br>**Metabolizer** | Reduced clopidogrel active metabolite formation; increased on-treatment platelet reactivity; increased risk for adverse cardiac and cerebrovascular events | Avoid standard dose (75 mg) clopidogrel if possible. Use prasugrel or ticagrelor at standard dose if no contraindication. | 0\|1 | 1 |
| **Clopidogrel**<br>**CYP2C19**<br>**Intermediate**<br>**Metabolizer** | NA | Results in lower systemic active metabolite concentrations, lower antiplatelet response, and may result in higher cardiovascular risk. Consider use of another platelet P2Y12 inhibitor. | 0\|1 | 1 |
| **Tacrolimus**<br>**CYP3A5**<br>**Normal**<br>**Metabolizer** | Lower dose-adjusted trough concentrations of tacrolimus and decreased chance of achieving target tacrolimus concentrations. | Increase starting dose 1.5 to 2 times recommended starting dose. Total starting dose should not exceed 0.3 mg/kg/day. Use therapeutic drug monitoring to guide dose adjustments. | 1\|0 | 1 |
| **Phenytoin**<br>**CYP2C9**<br>**Intermediate**<br>**Metabolizer** | Genetic variation reduces conversion of phenytoin to inactive metabolites. This increases the risk of side effects. | The loading dose does not need to be adjusted.<br>For the other doses, use 75% of the standard dose and assess the dose based on effect and serum concentration after 7-10 days | 2\|0 | 2 |
| **Fosphenytoin** | Reduced phenytoin metabolism; | For first dose, use typical | 3\|0 | 3 |
| **CYP2C9**<br>**Intermediate**<br>**Metabolizer** | Higher plasma concentrations will increase probability of toxicities. | initial or loading dose. For subsequent doses, use approximately 25% less than typical maintenance dose. Subsequent doses should be adjusted according to therapeutic drug monitoring, response and side effects. |  |  |
| **Tacrolimus**<br>**CYP3A5**<br>**Intermediate**<br>**Metabolizer** | Lower dose-adjusted trough concentrations of tacrolimus and decreased chance of achieving target tacrolimus concentrations. | Increase starting dose 1.5 to 2 times recommended starting dose. Total starting dose should not exceed 0.3 mg/kg/day. Use therapeutic drug monitoring to guide dose adjustments. | 3\|0 | 3 |

We identified four unique genes (*ABCG2*, *CACNA1S*, *CYP4F2*, *NUDT15*, eTable 22) that demonstrated potential utility for PGx clinical implementation from ES as they exhibited full coverage for every PGx phenotype identified. This is reflective of previous studies on the use of ES for PGx analysis which identified that approximately 93% of PGx variants with CPIC guidelines are covered by ES, including all or most of the variants located in the genes identified in our study.^56,57^ This suggests that for a subset of pharmacogenes, ES may be sufficient to support clinically meaningful PGx interpretation.

## Limitations

No outcome data were included. Therefore, we were unable to determine adverse drug event associations for individuals given medications for which PGx phenotype suggested deviation from standard of care. Prospective studies are needed to validate the clinical impact of PGx-guided medication therapy in pediatric critical care. ES is suboptimal to ascertain full PGx phenotypes although we demonstrate its general efficacy. The research ES were not held to the regulatory standards of a clinical laboratory. Future studies should investigate Genome-Sequencing or PGx targeted sequencing. Although our cohort was representative of an ancestrally diverse PICU, the small sample size and single-center nature of our study may limit the generalizability of our findings. Differences between the EMR-only and EMR+ES cohorts likely reflect several factors: ascertainment bias (EMR+ES patients were sequenced due to suspected genetic disease, while EMR-only captures all PICU admissions), different medication exposure windows, and different EMR systems with different prescribing protocols. These factors may limit direct comparisons between groups and affect generalizability.

## Conclusions

Our results demonstrate the potential clinical impact of PGx-guided dosing in a diverse PICU by revealing clinical recommendations to optimize medication dosing and avoid adverse medication events in this vulnerable population. This retrospective analysis found that many critically ill children receive medications with established PGx guidelines. As PGx guidelines are available for many commonly used medications but are not routinely implemented in clinical care, this represents a missed opportunity to increase efficacy and decrease toxicity. Using exome sequencing can effectively identify PGx phenotypes for a set of medications used in the pediatric intensive care unit although opportunities to identify some phenotypes are missed. The potential impact of PGx-guided prescribing may be greater in more medically complex patients, who are more likely to receive multiple medications with established PGx guidelines. These data lay the groundwork for further research and implementation efforts that are warranted to fully realize the benefits of PGx-guided precision medicine in pediatric critical care.

## Data Availability

All data produced in the present study are available upon reasonable request to the authors

## Acknowledgements

We acknowledge and miss the late Daniel Hughes for his contribution to alignments in this paper.

## Author contributions

Conceptualization: NL, ARP, SK, HB, NE, AGG, JEM Data curation: NL, ARP, CAB, JEM

Formal Analysis: NL, ARP, ASG, JEM Funding acquisition: JEM

Investigation: NL, ARP, ASG, JB, JBW, NE, JEM Methodology: NL, ARP, JEM

Project administration: NL, ARP, CAB, JEM Resources: JEM

Software: NL, JEM Supervision: JEM Visualization: NL, NE, JEM

Writing—original draft: NL, ARP, JEM

Writing—review & editing: NL, ARP, ASG, JB, JBW, NE, CB, SK, HB, AGG, JEM

