## Supplement for "Critically Ill Children Frequently Receive Medications with Established but Unused Pharmacogenomic Guidelines: Actionable Findings from an Integrated Electronic Medical Record and Exome Sequencing Study"

Lynch et al.

**eFIGURES**

**eTABLES**

### eFIGURES

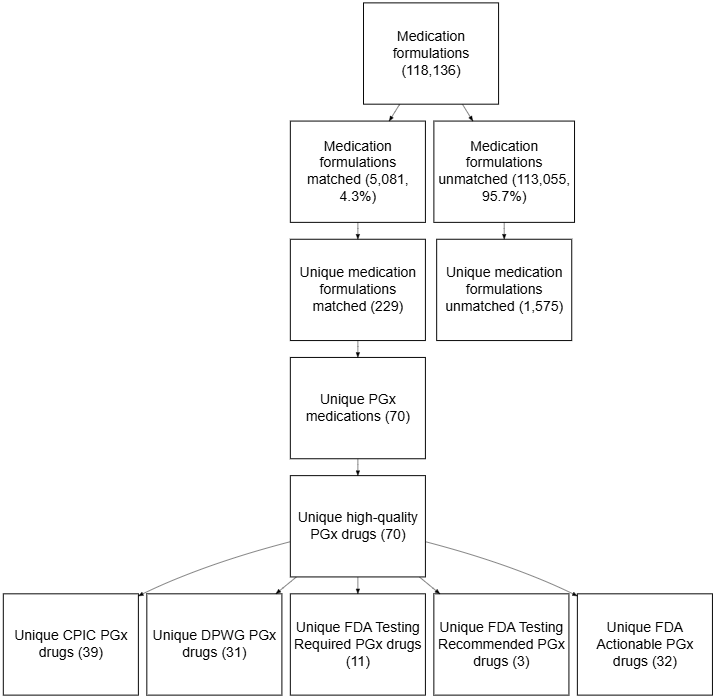

eFigure 1: Medication formulation matching in electronic medical record (EMR-only) cohort

Medication formulation matching in electronic medical record (EMR-only) cohort.Flowchart shows the matching of 118,136 medication formulations administered to the 4,939 PICU/CICU patients in the EMR-only cohort to medications with pharmacogenomic guidance. There were 74,232 unique patient/medication formulation combinations (median = 8 unique formulations per patient, range 1 - 196, IQR 15 - 18).

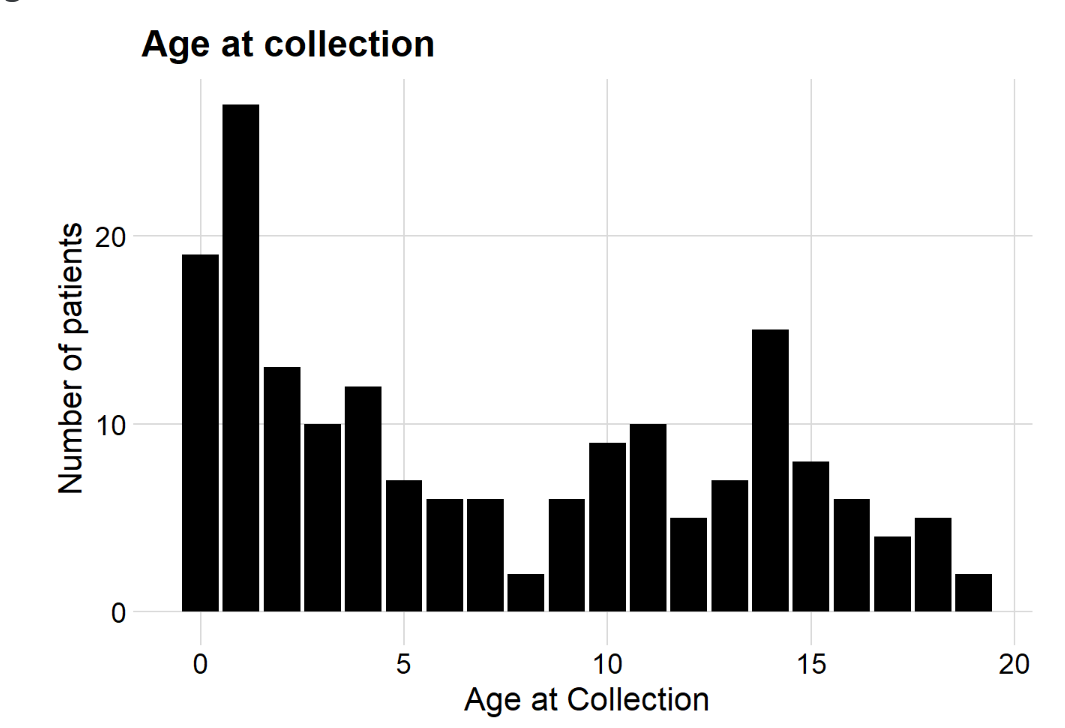

eFigure 2: Age at collection

Age at collection. Of the 192 individuals with both MAR and PGx, 179 have an age at collection. Age at collection (years) of exomes for children with both pharmacogenomic phenotypes, electronic medical record medication administration record data, and known age at collection (n = 179). Range 0.0 to 19.0, median = 5.8, interquartile range 1.3 - 13.0.^1^

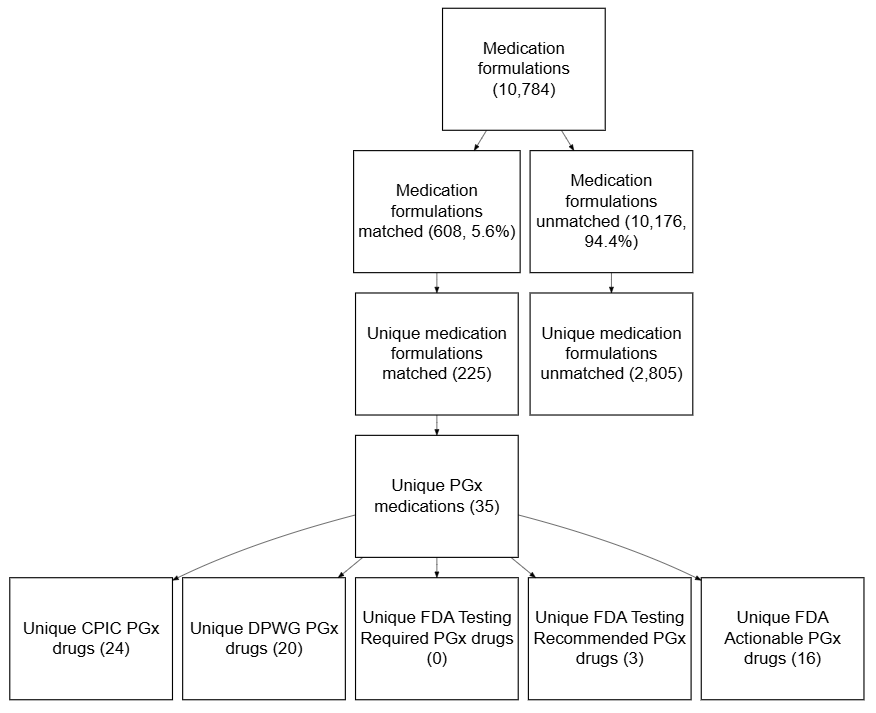

eFigure 3: Medication formulation matching in electronic medical record and exome sequencing (EMR + ES) cohort

Medication formulation matching in EMR + ES cohort. Flowchart shows the matching of 10,784 medication formulations administered to the 192 PICU/CICU patients in the EMR + ES cohort to medications with pharmacogenomic guidance. There were 10,784 unique patient/medication formulation combinations (median = 37 unique formulations per patient, range 1 - 367, IQR 56 - 80).

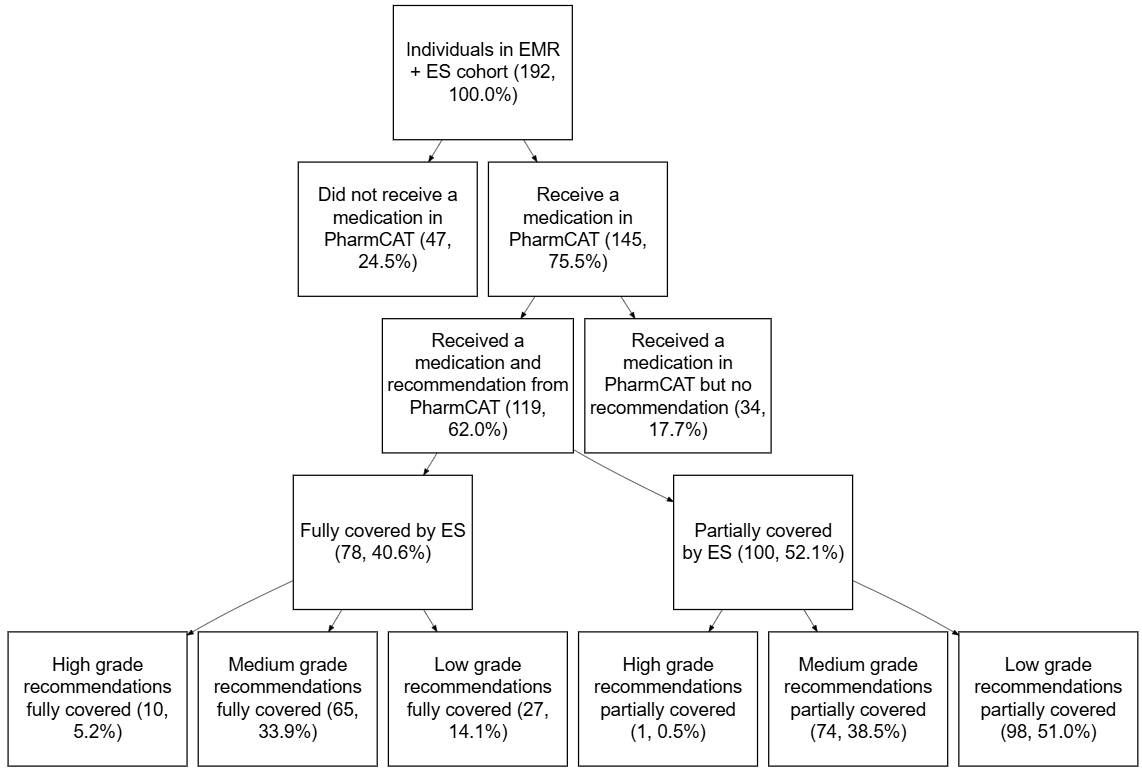

eFigure 4: Individuals in electronic medical record and exome sequencing (EMR + ES) cohort with recommendations by coverage and by grade

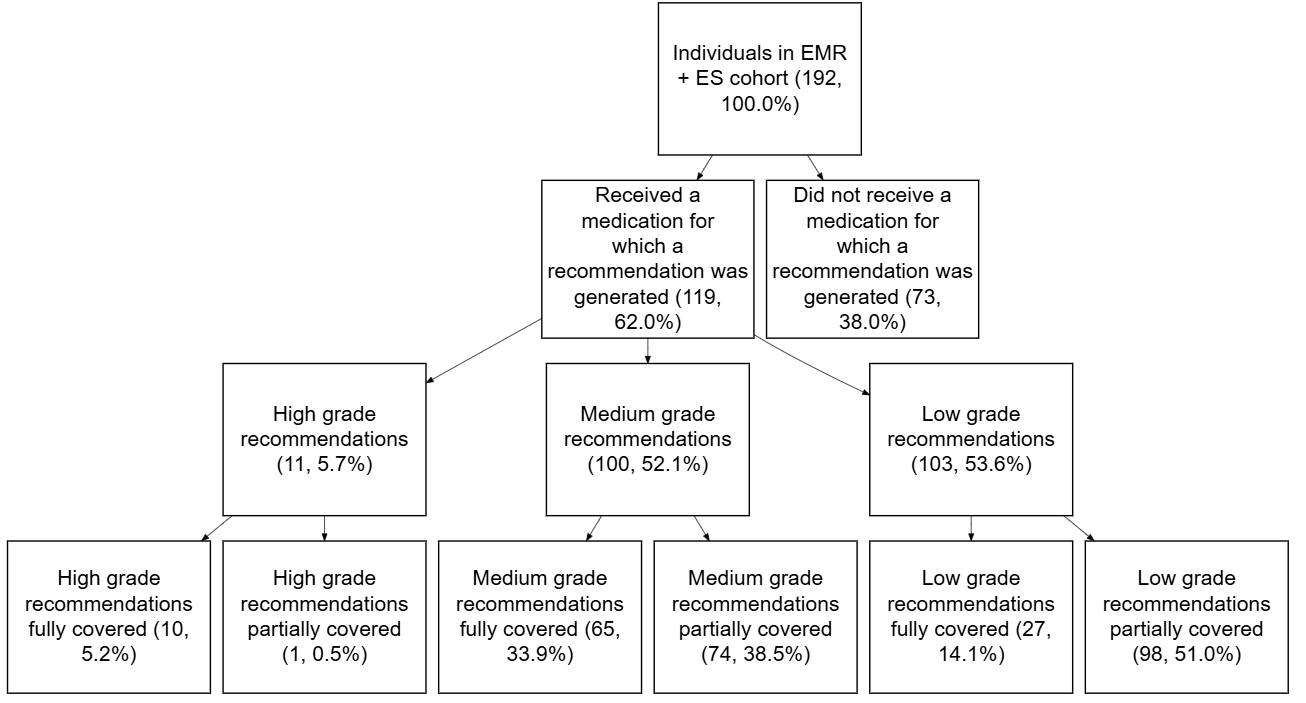

eFigure 5: Individuals in electronic medical record and exome sequencing (EMR + ES) cohort with recommendations

Flowchart chart showing individuals in EMR+ES cohort who received a medication with pharmacogenomic guidance. Groups are divided into individuals who received medications with pharmacogenomic guidance vs. those that did not. These categories are mutually exclusive. Of those who did receive a medication with pharmacogenomic guidance, groups are divided into those for whom a recommendation was generated from PharmCAT based on exome data vs. received a medication with pharmacogenomic guidance but did not receive a recommendation from PharmCAT. These groups are mutually exclusive. These groups are mutually exclusive. Among those who received a medication with pharmacogenomic guidance, groups are divided among those whose star diplotypes were fully covered by exome for at least one medication with pharmacogenomic guidance vs. only partially covered. These groups are mutually exclusive. Among recommendations generated by PharmCAT, individuals are grouped based on the degree of clinical change. These groups were not mutually exclusive.

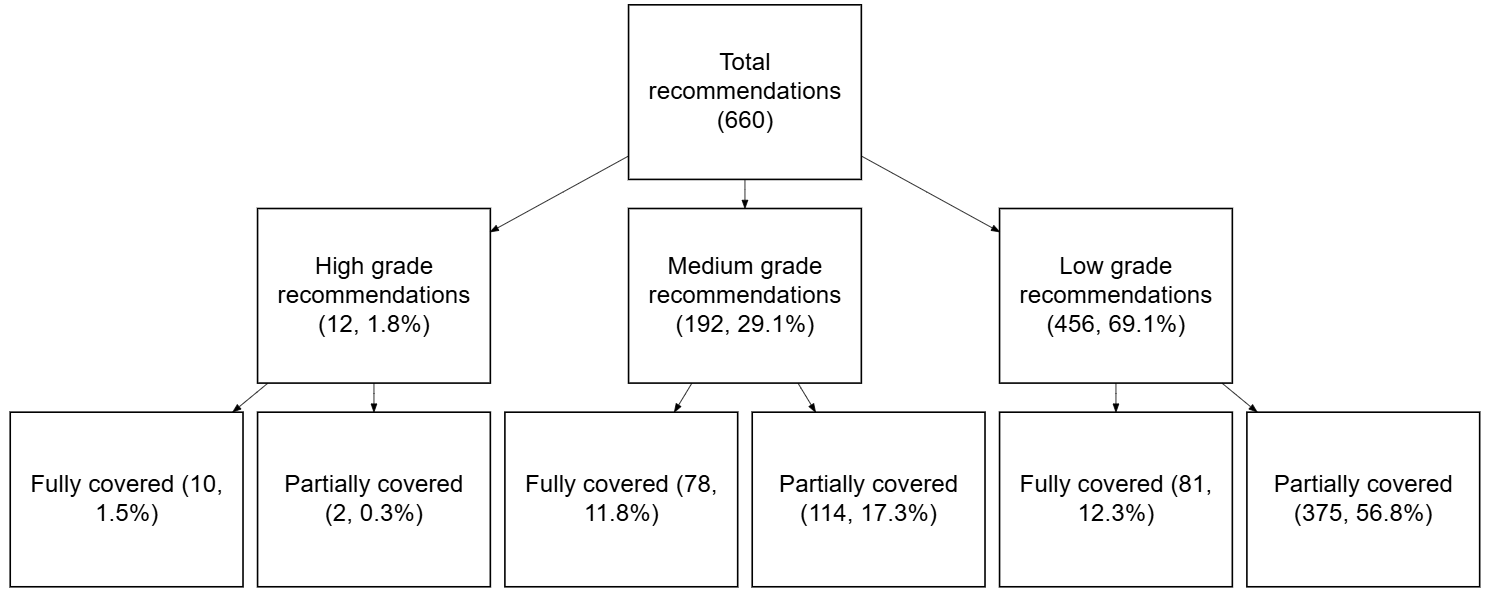

eFigure 6: Recommendations in electronic medical record and exome sequencing (EMR + ES) cohort

Flowchart chart showing recommendations in EMR+ES cohort who received a medication with pharmacogenomic guidance. Groups divided into recommendations with low, medium, or high grade clinical impact. These groups are mutually exclusive. Each recommendation clinical impact category divided into those recommendations generated from exome sequencing for which all positions were covered (“fully covered”) vs. some genomic positions were missing coverage (“partially covered”). These groups are mutually exclusive.

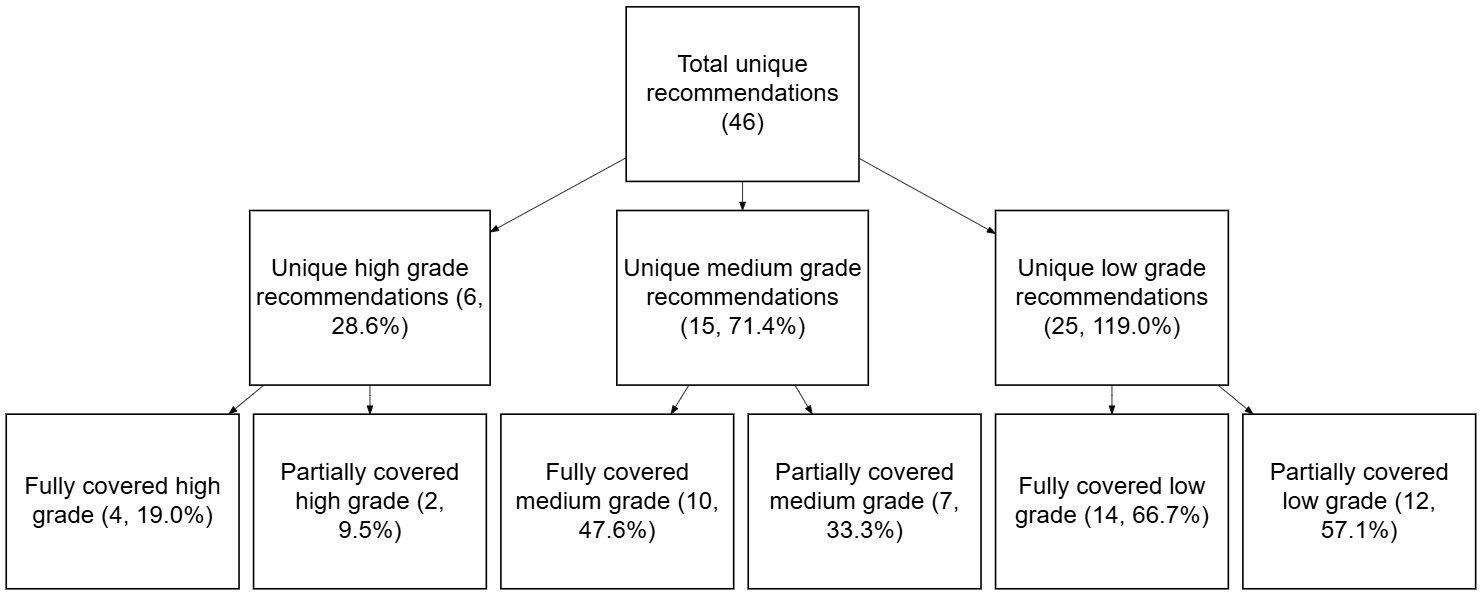

eFigure 7: Unique recommendations in electronic medical record and exome sequencing (EMR + ES) cohort

Flowchart chart showing individuals in EMR+ES cohort who received a medication with pharmacogenomic guidance. Groups divided into individuals who received medications for a pharmacogenomic phenotype could be fully discerned from exome sequencing (or not) and degree of clinical change indicated by the pharmacogenomic guidance.

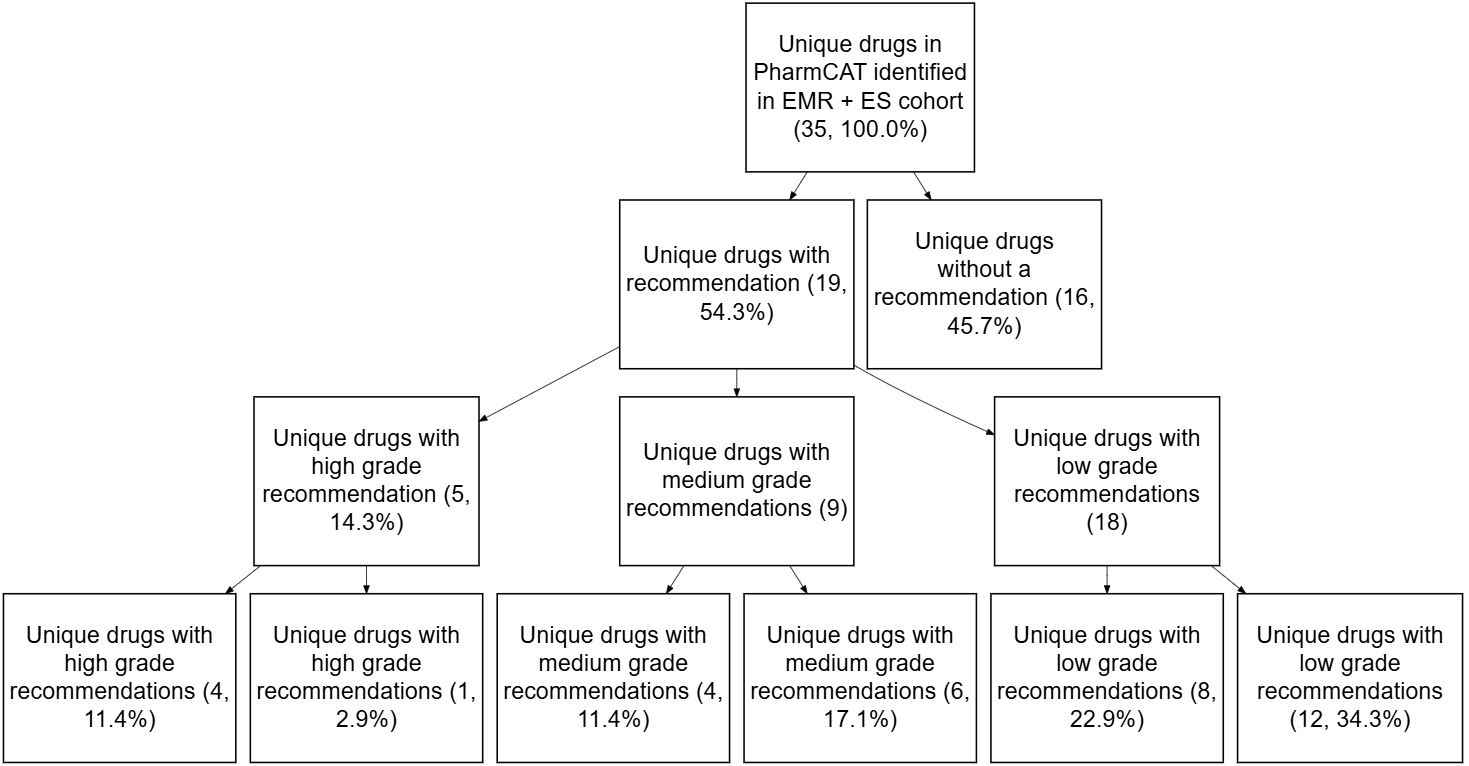

eFigure 8: Medications in electronic medical record and exome sequencing (EMR + ES) cohort with and without recommendations

Flowchart chart showing medications in EMR+ES cohort who received a medication with pharmacogenomic guidance. Groups divided into medications who received medications for a pharmacogenomic phenotype could be fully discerned from exome sequencing (or not) and degree of clinical change indicated by the pharmacogenomic guidance.

### eTABLES

eTable 1: Electronic medical record (EMR) actions indicating a medication has been given

EMR 1 was used prior to 2/1/2020 and EMR 2 was active from 2/1/2020 onward. These actions indicate that the medication was administered to the patient to filter out those orders entered into the EMR but never given to the patient. See excel.

eTable 2: Pharmacogenomic guidance by drug and type of guidance from CPIC and DPWG

Drugs and type of pharmacogenomic guidance from CPIC and DPWG. Guidance downloaded from The Pharmacogenomics Knowledgebase (PharmGKB) (downloaded 2/4/2025). CPIC = Clinical Pharmacogenetics Implementation Consortium, DPWG = Dutch Pharmacogenomics Working Group, Pediatric = carries ‘Pediatric’ tag in PharmGKB, Dose_Adj = if genotype is known, recommendation for a dose adjustment is possible, Alt_Drug = if genotype is known, recommendation for an alternative drug is possible, Other_Guidance = if genotype is known, a recommendation pertaining to neither dosing nor alternative drugs is possible, Testing_Info = recommendation for pharmacogenomic testing. See excel.

eTable 3: Genes with pharmacogenomic guidance from CPIC or DPWG

Genes with pharmacogenomic guidance from CPIC and DPWG. Guidance downloaded from The Pharmacogenomics Knowledgebase (PharmGKB) (downloaded 2/4/2025). CPIC = Clinical Pharmacogenetics Implementation Consortium, DPWG = Dutch Pharmacogenomics Working Group. See excel

eTable 4: Pharmacogenomic guidance by drug and type of guidance from the Food and Drug Administration (FDA)

Drugs and type of pharmacogenomic guidance from the FDA including only those with one of Testing Required, Testing Recommended, or Actionable PGx. Guidance downloaded from The Pharmacogenomics Knowledgebase (PharmGKB) (downloaded 2/4/2025). FDA = Food and drug administration. Testing Required = genetic or functional assay should be conducted before using this drug, Testing Recommended = genetic or functional assay is recommended before using this drug, Actionable PGx = if the patient’s genotype is known, a dose adjustment, contraindication or alternate drug recommendation, or other guidance is recommended although determining genotype/phenotype before using the drug is not necessarily recommended.

eTable 5: PGx medications eligible for matching to EMR medication formulations in EMR-only cohort

Table shows 272 medications with CPIC, DPWG, or clinically actionable FDA guidance, which were attempted to match to medication formulations in EMR-only cohort in EMR 2.

eTable 6: Medication formulations in EMR-only cohort matched to PGx medications in EMR 2

Formulations in EMR 2 administered to EMR-only cohort which were matched to PharmGKB medications with PGx guidance. See excel.

eTable 7: Medication formulations in EMR-only cohort unmatched to PGx medications in EMR 2

Formulations in EMR 2 administered to EMR-only cohort unmatched to PharmGKB medications with PGx guidance or excluded from matching. Formulations not administered systemically were identified with the following phrases (EX OINT, EX CREA, EX PATCH, EX GEL, OP OINT, OP SOLN, OP SUSP. IN NEBU) and excluded from matching. See excel.

eTable 8: Extreme phenotype identification and phenotype frequency estimates

Table shows choice of “extreme” phenotypes which indicate the largest clinical change for gene/drug combinations. Only medications administered to greater than or equal to 50 patients in the EMR-only cohort were considered. Table also shows estimated frequency of phenotype and includes citations used for estimating phenotype frequency. See excel.

eTable 9: Capture kits in EMR + ES cohort

Capture kits used in research exomes (n = 192). See Excel.

eTable 10: Genes for which PharmCAT issues guidance based on a VCF file

List of genes for which PharmCAT identifies star alleles, assigns phenotypes, and issues guidance based on a provided VCF. CPIC = Clinical Pharmacogenetics Implementation Consortium, DPWG = Dutch Pharmacogenomics Working Group. See excel

eTable 11: Phenotypes identified by PharmCAT

Phenotypes identified by PharmCAT. Includes phenotypes for genes requiring outside calls which are not relevant to this study. CPIC = Clinical Pharmacogenetics Implementation Consortium, DPWG = Dutch Pharmacogenomics Working Group. See excel

eTable 12: Medications for which PharmCAT issues guidance

183 unique medications for which PharmCAT issues guidance. PharmCAT issues guidance from CPIC, DPWG, FDA Labels and FDA PGx Associations (https://www.clinpgx.org/page/drugLabelLegend). Only FDA information for genes with CPIC or DPWG guidelines is issued because the FDA does not offer any genotype-to-phenotype mapping information. PharmCAT uses CPIC genotype-to-phenotype mappings when they exist, and DPWG genotype-to-phenotype mappings when no CPIC mappings exist, to determine the phenotypes to use with FDA label annotations and Table of Pharmacogenetic Associations (https://www.clinpgx.org/fdaPgxAssociations) entries. CPIC = Clinical Pharmacogenetics Implementation Consortium, DPWG = Dutch Pharmacogenomics Working Group, FDA = Food and Drug Administration. FDA PGx Association = https://www.fda.gov/medical-devices/precision-medicine/table-pharmacogenetic-associations. See excel

eTable 13: Medications in both PharmCAT and the clinically actionable CPIC/DPWG/FDA list

156 medications which were in both PharmCAT (182 medications, eTable 12) and the clinically actionable list of medications from PharmGKB with CPIC, DPWG, or clinically actionable FDA guidance (272 medications, eTable 5) and were used to identify pharmacogenomic medications in the EMR+ES cohort.

eTable 14: Medication formulations matched to PGx medications in EMR 1 from the EMR+ES cohort

Formulations in electronic medical record matched to PharmGKB medications analyzed in the EMR+ES cohort (eTable 5). See excel.

eTable 15: Medication formulations unmatched in EMR 1 from the EMR-only cohrt

Formulations in electronic medical record unmatched to pharmacogenomic medications (eTable 5) or excluded from matching. Formulations not administered systemically were identified with the following phrases (EX OINT, EX CREA, EX PATCH, EX GEL, OP OINT, OP SOLN, OP SUSP. IN NEBU) and excluded from matching. See excel.

eTable 16: Medications in the clinically actionable CPIC/DPWG/FDA list from the EMR-only cohort for which PharmCAT does not offer recommendations.

Of the 272 pharmacogenomic medications (eTable 5) from PharmGKB with CPIC, DPWG, or clinically actionable FDA guidance, 116 medications are not issued guidance by PharmCAT.

eTable 17: Pharmacogenomic medications per PICU patient in EMR-only cohort

See legend for Figure 2 for details. See Excel

eTable 18: Pharmacogenomic medications per PICU patient in EMR-only cohort

Expanded list from Table 1 showing all medications administered to critically ill children. CPIC = Clinical Pharmacogenetics Implementation Consortium, DPWG = Dutch Pharmacogenomics Working Group, FDA = Food and Drug Administration, Pediatric = carries ‘Pediatric’ tag in PharmGKB, Dosing = if genotype is known, recommendation for a dose adjustment is possible, Alternate Drug = if genotype is known, recommendation for an alternative drug is possible, Testing = guideline contains information about pharmacogenetic testing, Other = if genotype is known, a recommendation pertaining to neither dosing nor alternative drugs is possible, Testing Info = recommendation for pharmacogenomic testing, FDA Testing Required = genetic or functional assay should be conducted before using this drug, FDA Testing Recommended = genetic or functional assay is recommended before using this drug, FDA Actionable PGx = if the patient’s genotype is known, a dose adjustment, contraindication or alternate drug recommendation, or other guidance is recommended although determining genotype/phenotype before using the drug is not necessarily recommended. See excel.

| **Gene** | **Unique Patients (% Cohort)** |
| --- | --- |
| *CYP2C9* | 853 (17.3%) |
| *CYP2C19* | 633 (12.8%) |
| *CYP2D6* | 462 (9.4%) |
| *CYP3A5* | 182 (3.7%) |
| *HLA-B* | 167 (3.4%) |
| *CYP4F2* | 107 (2.2%) |
| *VKORC1* | 107 (2.2%) |
| *CYP3A4* | 52 (1.1%) |
| *CYP2B6* | 45 (0.9%) |
| *G6PD* | 41 (0.8%) |
| *ABCG2* | 19 (0.4%) |
| *NUDT15* | 17 (0.3%) |
| *TPMT* | 17 (0.3%) |
| *SLCO1B1* | 15 (0.3%) |
| *MT-RNR1* | 9 (0.2%) |
| *HLA-A* | 7 (0.1%) |
| *CACNA1S* | 6 (0.1%) |
| *RYR1* | 6 (0.1%) |
| *DPYD* | 1 (0.0%) |
| *IFNL3* | 1 (0.0%) |
| *CFTR* | 0 (0.0%) |
| *UGT1A1* | 0 (0.0%) |

eTable 19: All pharmacogenes implicated in medications administered to EMR cohort

Table summarizing percentage of EMR-only cohort (4,939 individuals) given drugs for which the listed pharmacogenes contributes to the pharmacogenomic phenotype.

eTable 20: EMR+ES cohort taking pharmacogenomic medications by medication

Percentage of EMR+ES cohort taking medications with pharmacogenomic guidance issued by PharmCAT (eTable 13). See Excel.

eTable 21: Assessing efficacy of exome to determine pharmacogenomic phenotype

Drugs with adequate coverage for 192 individuals in EMR+ES cohort. The first row of the table can be interpreted as 135 individuals (of the 192) had adequate coverage in their exome sequencing for all positions in the star alleles of 40 medications (of the 105 medications for which PharmCAT issues CPIC or DPWG guidance, eTable 2). See Excel.

eTable 22: Effective star allele identification using exomes by drug and capture kit

Summary of individuals and medications for which all positions of star alleles were adequately covered by capture kit. “Medication in EMR” indicates that the drug was administered to at least one individual in the EMR+ES cohort. Medications limited to those with CPIC or DPWG guidance (eTable 2). See Excel.

eTable 23: Pharmacogenes with adequate coverage in EMR+ES cohort by capture kit.

Summary of number of pharmacogenes for which all star alleles were adequately covered by capture kit in EMR+ES cohort. Limited to definitions from CPIC and DPWG. See Excel.

eTable 24: Pharmacogene star allele positions and coverage by exome capture kit

Number of individuals with adequate coverage at each star allele position by exome capture kit. Positions defined by CPIC and DPWG. See Excel.

eTable 25: Pharmacogenomic recommendations from PharmCAT for EMR+ES cohort

All recommendations issued by PharmCAT for EMR+ES cohort. Recommendation text may be abbreviated for display purposes. ‘Covered|Not covered’ indicates the number of individuals with the phenotypes for which all star allele positions are covered by exome or not. CPIC = Clinical Pharmacogenetics Implementation Consortium, DPWG = Dutch Pharmacogenomics Working Group, ᵃ indicates CPIC guideline, ᵇ indicates DPWG guideline, ᶜ indicates FDA label, ᵈ indicates FDA Association. Dose change indicates how much a recommendation would change medication administration (1 indicates significant change, 2 indicates a mild change, 3 indicates no change). Total recs indicates the number of individuals in the EMR+ES cohort receiving this recommendation. See Excel.

| Ancestry | Percent cohort |
| --- | --- |
| South Asian | 5.7 |
| Middle Eastern | 2.1 |
| Latino | 33.9 |
| European | 16.7 |
| East Asian | 4.7 |
| African | 29.7 |
| Admixed | 7.3 |

eTable 26: Geographic ancestry of EMR+ES

EMR+ES cohort includes 192 individuals. Geographic ancestry was calculated using a pre-trained neural-network generated probability estimates for each of six groups (European, African, Latino, East Asian, South Asian and Middle Eastern).
